# Antipsychotics Lower Peripheral Markers of Inflammation in Drug-naïve Early Psychosis: A Pilot Study

**DOI:** 10.1101/2025.08.07.25333214

**Authors:** Nicole Šafářová, Marián Kolenič, Ivana Tašková, Václav Čapek, Petra Fürstová, Filip Španiel

**Affiliations:** National Institute of Mental Health, Klecany, Czech Republic; 3rd Faculty of Medicine at Charles University, Prague, Czech Republic; Psychiatric Hospital Bohnice, Prague, Czech Republic; Faculty of Pharmacy at Charles University, Hradec Králové, Czech Republic

**Keywords:** neutrophil-to-lymphocyte ratio, monocyte-to-lymphocyte ratio, platelet-to-lymphocyte ratio, systemic immune-inflammation index, neuroinflammation, early psychosis, first-episode schizophrenia, antipsychotic medication, cumulative chlorpromazine equivalents

## Abstract

**Introduction:** Neuroinflammation is increasingly recognized as a core pathophysiological mechanism in schizophrenia and can be indirectly assessed through peripheral inflammatory markers. Therefore, this pilot study investigated the impact of antipsychotic treatment on inflammation in patients with first-episode psychosis (FEP) who were antipsychotic-naive at study entry.

**Methods:** Thirty-three drug-naïve FEP patients provided blood samples upon admission (V0) and follow-up (V1), from which peripheral inflammatory markers—i.e., neutrophil/lymphocyte ratio (NLR), monocyte/lymphocyte ratio (MLR), platelet/lymphocyte ratio (PLR), and systemic immune-inflammation index (SII)—were calculated. Antipsychotic doses during continuous hospitalization between V0 and V1 (34 days, IQR: 21-49 days) were converted into cumulative chlorpromazine equivalents (cCPZ).

**Results:** In multiple regression models adjusting for sex, age, BMI, DUP, and clozapine use, cumulative antipsychotic exposure significantly predicted reductions in ΔNLR (p = 0.048), ΔMLR (p = 0.041), ΔPLR (p = 0.028), and ΔSII (p = 0.028). All associations remained significant following false discovery rate adjustment (pFDR = 0.048 for all outcomes).

**Conclusion:** These findings suggest a consistent dose-dependent anti-inflammatory effect during early antipsychotic treatment in FEP. Given the exploratory nature of this study, larger studies are needed to confirm these findings.

## 1. Introduction

Schizophrenia is a severe mental illness affecting ∼1% of the global population (Amelang 2023) with a significant functional impairment and great disability (Charlson et al. 2018; Vos 1990). Traditionally, schizophrenia’s pathophysiology has been connected to the dopamine hypothesis, which suggests that hyperactivity of dopaminergic transmission in subcortical regions—particularly the mesolimbic pathway—underlies positive symptoms such as hallucinations and delusions (Toda and Abi-Dargham 2007). This theory guides current treatment centered on dopamine-2-receptor antagonism (Seeman 2021), which, while effective on positive symptoms, is largely ineffective in managing negative symptoms and cognitive decline (Marder 1999)—key determinants of functional outcomes. As a result, current treatment alleviates only a fraction of the total disease burden (Andrews et al. 2003; Fervaha et al. 2015).

In this context, the neuroinflammatory pathway offers a compelling framework for understanding the pathophysiology of schizophrenia. Similar to neurodegenerative diseases, schizophrenia is associated with neurodegenerative changes (Stone et al. 2022; Gupta and Kulhara 2010) linked to chronic, low-grade inflammation and may potentially result in symptom progression (Howes and McCutcheon 2017; Khandaker and Dantzer 2016). In this neurodegeneration-inflammation framework, diverse neurodegenerative processes induce cellular stress that releases damage-associated molecular patterns (DAMPs) (Castro-Gomez and Heneka 2024). DAMPs activate pattern recognition receptors—such as Toll-like receptors (TLRs) on glial cells—initiating neuroinflammatory cascades marked not only by the central release but also the peripheral spillover of cytokines, including IL-1β, IL-6, TNF-α, and chemokines CXCL8 (IL-8), CCL2 (MCP-1), and CXCL10 (IP-10). (Olson and Miller 2004; Sokol and Luster 2015). This cascade is increasingly recognized as a pathophysiological hallmark of neuropsychiatric disorders, including schizophrenia. (Singh 2022; Müller 2019; Ermakov et al. 2023; Ransohoff and Brown 2012).

Chemokines drive the recruitment and activation of neutrophils, monocytes, and lymphocytes, linking central neuroinflammation to peripheral immune responses (Sokol and Luster 2015). This connection is reflected in elevated peripheral inflammatory markers—such as NLR, MLR, PLR, and SII—consistently reported in schizophrenia (Mazza et al. 2020; Karageorgiou et al. 2019; Sandberg et al. 2021). Such immune alterations are evident from the earliest stages of psychotic illnesses (Trovão et al. 2019)—individuals at clinical high risk (CHR) and with first-episode psychosis (FEP) show increased inflammatory markers, including IL-6 and NLR (Misiak et al. 2021; Dunleavy et al. 2022; Sandberg et al. 2021; Karageorgiou et al. 2019; Capuzzi et al. 2017; Zhou et al. 2021). These changes may contribute to disease progression and greater disease severity (Perkins et al. 2015; Dunleavy et al. 2022; Zhou et al. 2020; Wang et al. 2024; Šafářová et al. 2025).

Although most second-generation antipsychotics (APs) exhibit anti-inflammatory action through the reduction of cytokines and chemokines in FEP (Capuzzi et al. 2017; Marcinowicz et al. 2021; Šafářová et al. 2025)—their impact on peripheral markers remains equivocal (Fernandes et al. 2016; Sandberg et al. 2021; Šafářová et al. 2025).

Given these uncertainties, a more precise understanding of how AP treatment influences neuroinflammation in early psychosis is needed. Therefore, the present study aimed to evaluate whether cumulative AP treatment is associated with changes in NLR, MLR, PLR, and SII in drug-naïve FEP patients. To our knowledge, the present study is the first to explore the anti-inflammatory action of APs in FEP using cumulative chlorpromazine dose equivalents (cCPZ) as an assessment of drug exposure across diverse regimens while controlling for confounders such as age, sex, clozapine use, and duration of untreated psychosis (DUP). Moreover, we examined a comprehensive panel of hemato-immunological markers—including NLR, MLR, PLR, and SII—to more fully capture the peripheral immune response to antipsychotic treatment.

## 2. Methods

### 2.1. Study setting

This retrospective pilot study analyzed data from the ongoing ESO project at the National Institute of Mental Health, Klecany, Czech Republic.

Initial drug-naive blood samples (V0) were collected at the Psychiatric Hospital Bohnice in Prague, with follow-up assessments (V1) conducted at the National Institute of Mental Health in Klecany. All patients included in this study provided informed consent.

Data for this study were extracted from the ESO database in January 2023. All participants provided written informed consent, and the study was conducted in accordance with the latest version of the Declaration of Helsinki. The study protocol was reviewed and approved by the Research Ethics Board of the National Institute of Mental Health in Klecany (Approval No. 127/17).

### 2.2. Inclusion criteria

- Drug-naïve patients with the first-episode psychosis (FEP)
- Continuous hospitalization between V0 and V1
- Medication and daily doses had to be consistently monitored throughout their hospitalization.
- No ongoing viral or bacterial infections between V0 and V1

These criteria led to the inclusion of 33 drug-naïve FEP patients in this study.

### 2.3. Data collection

Continuous hospitalization allowed for consistent monitoring of medication adherence. During the study period, administration of antipsychotic medication alongside dosage information was recorded daily. All doses were converted into chlorpromazine equivalents, and cumulative doses were calculated for each patient to facilitate comparison across all of the study participants.

Blood samples were collected at two predefined time points—V0 and V1. Standard complete blood counts (CBCs) were provided, including the absolute count of neutrophils, lymphocytes, monocytes, and platelets. Hemato-immunological markers, including NLR, MLR, PLR, and SII, were computed using the following formulas:

- NLR = neutrophils/lymphocytes
- MLR = monocytes/lymphocytes
- PLR = platelets/lymphocytes
- SII = (platelets × neutrophils)/lymphocytes

For each marker, the change between the two time points (Δ = V1 − V0) was calculated and used as the dependent variable in the subsequent analysis. The SII was included because it reflects a combined index of neutrophil, lymphocyte, and platelet counts, offering a broader and potentially more sensitive measure of systemic inflammation relevant to psychotic disorders. Higher values of NLR, MLR, PLR, and SII are generally interpreted as indicators of elevated peripheral inflammation. Therefore, in this study, a decrease in these markers is suggestive of a reduction in neuroinflammatory activity.

### 2.4. Covariate selection

Cumulative antipsychotic exposure (cCPZ) was the prespecified primary predictor of interest. All other variables were included to account for their potential confounding effects on inflammation. Specifically, **sex** was included due to the known sex differences in immune response, with males typically exhibiting higher basal inflammation (Klein and Flanagan 2016; Martínez de Toda et al. 2023). **Age** was included given its established association with increased inflammation (Singh et al. 2024; Li et al. 2023). Although the evidence for duration of untreated psychosis (**DUP**) is inconclusive, it was included due to its possible association with inflammation in early psychosis (Prakash et al. 2025; Kim et al. 2023; Zoghbi et al. 2023). **Clozapine** use was included to account for the potential confounding effect of clozapine-induced neutropenia (CIN) and its other known alterations of white blood cells. Finally, body mass index (**BMI**) was included to control for the metabolic contribution to low-grade inflammation (Su et al. 2024).

### 2.5. Data analysis

Associations between cumulative antipsychotic exposure and changes in inflammatory markers (ΔNLR, ΔMLR, ΔPLR, ΔSII) were examined using multiple regression models including sex, age, BMI, duration of untreated psychosis (DUP), and clozapine use as covariates. Regression assumptions were evaluated visually using residual diagnostics. False discovery rate (FDR) adjustment (Benjamini–Hochberg) was applied across the four outcomes to account for multiple testing. Bonferroni-adjusted *p*-values are reported in *Supplementary Table 1* for transparency but were not used to guide inference due to correlated endpoints and the exploratory nature of this pilot study.

All statistical analyses were performed using the R software (version 4.2.1). The dataset was screened for outliers and missing values; no patients were excluded, and no imputation was performed. Analyses included only complete cases. Statistical significance was set at *p* < 0.05 (two-tailed).

### 2.6. Power and sample size

This study was designed as an exploratory pilot to evaluate feasibility and estimate effect sizes for future research; therefore, no a priori power analysis was conducted. However, post hoc sensitivity and power analyses were done in the R software. With a total sample size of N = 33 and 6 predictors (cCPZ, sex, age, DUP, BMI, and clozapine use), this study had 80% power (α = 0.05) to detect large effects of approximately **Cohen’s f² = 0.521** (R² = 0.343) for the predictor set. Therefore, smaller or moderate effects may have gone undetected due to the limited power.

## 3. Results

### 3.1. Cohort analysis

The analysis included 33 drug-naïve FEP patients, of whom 11 were female and 22 were male. The mean age was 24.58 ± 5.43 years. The median time between V0 and V1 was 34 days (IQR: 21-49 days) . For more demographic information, see *Table 1*.

**Table 1:** Descriptive statistics for demographic and clinical variables (N = 33)

| | Mean ± SD | Median [IQR] |
| --- | --- | --- |
| DUP (days) |  | 27.00 [14.00, 116.00] |
| age at V0 (years) | $24.58 \pm 5.43$ | |
| Length of V0 to V1 (days) |  | 34.00 [21.00, 49.00] |
| Length of AP treatment to V0 (days) | $6.33 \pm 12.75$ | |
| BMI | $21.94 \pm 2.85$ | |

*V0—the initial visit, V1—the follow-up visit, DUP—the duration of untreated psychosis, AP—antipsychotic*

**Table 2:** Descriptive statistics for inflammatory markers at baseline (V0) and follow-up (V1)

| Marker | Mean V0 (SD) | Mean V1 (SD) | Mean $\Delta$ | p-value |
| --- | --- | --- | --- | --- |
| NLR | 1.89 (1.03) | 2.02 (1.23) | 0.13 | 0.576 |
| MLR | 0.228 (0.0831) | 0.245 (0.114) | 0.017 | 0.395 |
| PLR | 105 (31.1) | 130 (41.8) | 24.4 | **0.002\*\*** |
| SII | 421 (282) | 453 (293) | 31.6 | 0.605 |

**\*\* statistical significance $p < 0.01$**
*NLR—neutrophil-to-lymphocyte ratio, MLR—monocyte-to-lymphocyte ratio, PLR—platelet-to-lymphocyte ratio, SII—systemic immune-inflammation index, V0—the initial visit, V1—the follow-up visit*

Descriptive statistics for inflammatory markers at V0 and V1 are depicted in *Table 2*. The overall changes in NLR (mean Δ = 0.13, *p* = 0.576), MLR (mean Δ = 0.017, *p* = 0.395), and SII (mean Δ = 31.6, *p* = 0.605) were minimal and not statistically significant. Only PLR showed a significant increase (mean Δ = 24.4, *p* = 0.002), likely influenced by the high variability across patients (Δ range: –48.6 to +170.0).

The median cumulative antipsychotic exposure expressed in chlorpromazine equivalents (cCPZ) in the whole cohort was 16331 mg (IQR: 8114–46076 mg), while the mean cCPZ was 28370.1 mg. The majority of patients were prescribed olanzapine (81.8%, n = 27) and risperidone (78.8%, n = 26). Aripiprazole was used in 30.3% of patients (n = 10), clozapine and haloperidol both in 24.2% (n = 8), amisulpride and levomepromazine in 18.2% (n = 6), paliperidone in 9.1% (n = 3), ziprasidone in 6.1% (n = 2), and zuclopenthixol in 3.0% (n = 1). Throughout the entire study period (V0 to V1), 42.4% of patients (n = 14) were treated with antipsychotic monotherapy. The distribution of antipsychotic use is depicted in *Figure 1*, highlighting predominant use of olanzapine and risperidone among the patients, both of which are recommended as first-line treatments in psychiatric guidelines (Keepers et al. 2020) for managing acute psychosis and schizophrenia.

**Figure 1:**
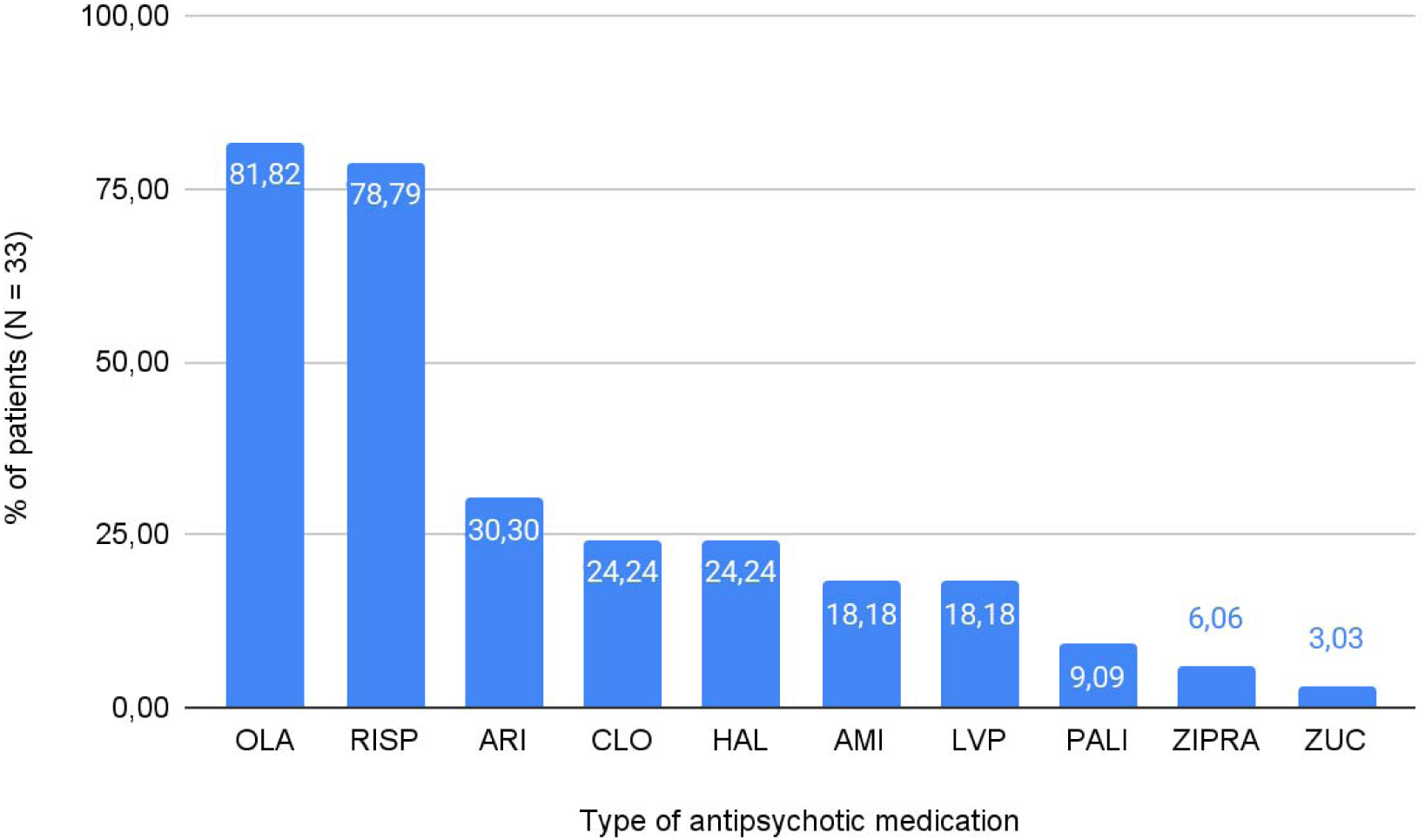
Distribution of antipsychotic use in patient cohort (N = 33) OLA-olanzapine, RISP-risperidone, ARI-aripiprazole, CLO-clozapine, HAL-haloperidol, AMI-amisulpride, LVP-levomepromazine, PALI-paliperidone, ZIPRA-ziprasidone, ZUC-zuclopenthixol

### 3.2. Association between cumulative antipsychotic exposure and inflammation

In multiple regression models with Δ inflammatory markers as outcomes (Δmarker ∼ cumulative CPZ-equivalents + sex + age + BMI + DUP + clozapine use), cumulative antipsychotic exposure significantly predicted reductions in ΔNLR (β = −2.22×10⁻⁵, 95% CI: −4.31×10⁻⁵ to −1.19×10⁻⁶, ***p* = 0.048**), ΔMLR (β = −1.71×10⁻⁶, 95% CI: −3.26×10⁻⁶ to −1.52×10⁻⁷, ***p* = 0.041**), ΔPLR (β = −7.83×10⁻⁴, 95% CI: −1.44×10⁻³ to −1.25×10⁻⁴, ***p* = 0.028**), and ΔSII (β = −6.47×10⁻³, 95% CI: −1.19×10⁻² to −1.04×10⁻³, ***p* = 0.028**). All associations remained statistically significant following FDR adjustment (***p*FDR = 0.048** for all outcomes). Full regression results are presented in *Table 3*.

To improve interpretability, regression coefficients were also expressed per +10,000 mg-days CPZ-equivalent exposure. On this scale, an additional +10,000 mg-days was associated with estimated reductions of −0.22 in ΔNLR, −0.02 in ΔMLR, −7.83 in ΔPLR, and −64.66 in ΔSII, indicating largest cCPZ–driven reductions in SII. Additionally, sex significantly predicted ΔMLR (***p* = 0.045**; ***p*FDR = 0.049**), with males showing lower reductions compared to females. Age, BMI, DUP, and clozapine use were not statistically significant predictors for any inflammatory marker.

**Table 3:** Adjusted regression models predicting change in inflammatory markers.

| Δ-marker | $\beta$ | 95% CI | $p$ | $pFDR$ |
| --- | --- | --- | --- | --- |
| NLR | -2.22e-05 | [-4.31e-05, -1.19e-06] | 0.0484* | 0.0484* |
| MLR | -1.71e-06 | [-3.26e-06, -0.15e-06] | 0.0409* | 0.0484* |
| PLR | -7.83e-04 | [-1.44e-03, -1.25e-04] | 0.0277* | 0.0484* |
| SII | -6.47e-03 | [-1.19e-02, -1.04e-03] | 0.0276* | 0.0484* |

\* statistical significance $p < 0.05$
*Cumulative exposure: unscaled $\beta$ and $p$ -values. Covariates: sex, age, BMI, DUP, and clozapine use.*
*NLR—neutrophil-to-lymphocyte ratio, MLR—monocyte-to-lymphocyte ratio, PLR—platelet-to-lymphocyte ratio, SII—systemic immune-inflammation index,*

## Discussion

This pilot study provides a unique perspective on the intricate relationship between antipsychotic therapy and inflammation in patients with FEP. By examining within-subject changes in inflammatory markers throughout their treatment, we extend previous research by moving beyond static, cross-sectional comparisons to a more dynamic assessment of immunological shifts during antipsychotic therapy. Our findings indicate that cumulative chlorpromazine equivalent doses were associated with reductions in peripheral inflammation markers (NLR, MLR, PLR, and SII), suggesting a potential dose-dependent anti-inflammatory effect of APs. Although the associations survived FDR correction, the effect estimates were imprecise, consistent with an exploratory pilot sample

Our findings of dose-dependent reductions in NLR, MLR, PLR, and SII following antipsychotic treatment in initially drug-naive patients may reflect a reduction in peripheral immune activation, which could represent a downstream component in the pathophysiological cascade of psychosis. One plausible mechanism by which antipsychotics may modulate inflammatory pathways is the modulation of the TLR-4/NF-κB cascade, through which antipsychotics may suppress glial activation and cytokine release (Matejuk and Ransohoff 2020; de Bartolomeis et al. 2022; Balaji et al. 2020; Weickert et al. 2024). In parallel, their potential anti-inflammatory action may involve dopaminergic modulation—particularly via D3 receptor-mediated astrocyte activation (Montoya et al. 2019; Pacheco 2017; Vidal and Pacheco 2020; Feng and Lu 2021) and subsequent NF-κB signalling.

Although the potential anti-inflammatory effects of antipsychotics may vary, particularly between the first- and second-generation agents—with evidence suggesting stronger anti-inflammatory action for SGAs (Capuzzi et al. 2017; Patlola et al. 2023; Šafářová et al. 2025)—we did not conduct a comparative analysis due to limited statistical power. Nonetheless, the observed dose-dependent effect based on cumulative chlorpromazine-equivalent dosing supports, at present, a general anti-inflammatory role for antipsychotics as a class, warranting further investigation.

Despite the promising findings, our study has several limitations. The small sample size (N = 33) limits the statistical power and generalizability—our study was only powered to detect large effects (f² ≈ 0.52) with 80% power. As such, our findings should be interpreted with caution. The retrospective design introduces potential biases related to timing, unmeasured confounders, and variability in clinical or laboratory data. Although we adjusted for key covariates (age, sex, DUP, BMI, and clozapine use) and excluded patients with acute viral or bacterial infections, other factors—such as comorbidities or somatic medications—could not be fully controlled for. These limitations call for more rigorous prospective research on larger cohorts. A further limitation is the treatment heterogeneity; all antipsychotics were grouped into a single cCPZ variable, regardless of their classification as either first-generation (FGA) or second-generation (SGA) subgroups. This approach facilitated standardized dose comparisons; however, it likely masked potential class-specific immunomodulatory effects, as previously noted (SGAs vs. FGAs). Future studies should stratify analyses by antipsychotic class to examine these differences more accurately. Another limitation concerns the interrelated nature of the inflammatory markers examined. The SII is mathematically derived from platelet, neutrophil, and lymphocyte counts, and therefore partly reflects the same signal captured by NLR and PLR. This inherent overlap may inflate associations or limit the ability to interpret each marker as an independent indicator of immune activity. Nonetheless, we included SII because it has been increasingly recognized as an integrative marker of systemic inflammation, even though most psychosis research to date has focused more extensively on NLR and related ratios. Finally, although all results survived FDR correction, they did not remain significant under the more conservative Bonferroni adjustment, underscoring the exploratory nature of the findings. Due to these limitations, our findings should be viewed as an initial signal that requires confirmation rather than definitive evidence of antipsychotic-mediated immunomodulation.

Clinically, inflammatory markers, such as the NLR, MLR, PLR, and SII, are appealing due to their low cost and broad availability, although their predictive utility has not yet been established. If validated in larger, prospective cohorts, these markers could serve as accessible and cost-effective tools for early detection, prognostication, and personalized intervention—particularly in identifying individuals at high risk for psychosis or poor treatment response. They could also help in detecting subclinical relapses and guide personalized antipsychotic strategies. Elevated baseline levels of these hemato-immunological markers might be indicative of a heightened risk for the development of psychosis. Therefore, future work may clarify whether early changes in these markers could inform intervention strategies. Importantly, our findings underscore the need to further explore the anti-inflammatory potential of APs. By focusing on their immunomodulatory properties, we may be able to develop novel approaches and strategies that could potentially treat the refractory symptoms of schizophrenia.

Future research must focus on prospective, longitudinal studies to evaluate the response of inflammatory markers to treatment and their potential to predict long-term outcomes in schizophrenia, including symptom persistence, relapse, and alterations in brain structure. Integrating neuroimaging data may elucidate the relationship between sustained neuroinflammation and progressive brain structural changes, including grey matter loss and altered connectivity, thereby offering insights into the neuroimmune mechanisms underlying schizophrenia progression. These studies would enhance the predictive validity of these biomarkers and facilitate their wider application in psychiatric practice.

## Conclusion

Our findings suggest that changes in hemato-immunological markers, such as NLR, MLR, PLR, and SII, may reflect neuroimmune modulation in response to antipsychotic treatment in FEP. Significant associations between cCPZ and ΔNLR, ΔMLR, ΔPLR, and ΔSII—even after controlling for age, sex, DUP, BMI and clozapine use—support the hypothesis that current antipsychotics may exert anti-inflammatory effects. Based on the knowledge from previous literature, these changes could be caused by the potential underlying antipsychotic’s mechanism of action. Our results align with the emerging view that antipsychotics may have a dual role in psychosis treatment—beyond their neurotransmitter effects, they could modulate inflammation, offering new insights into the pathophysiology and management of schizophrenia. While our study is limited by its small sample size and retrospective design, the findings contribute to a growing body of evidence supporting the role of neuroinflammatory processes in schizophrenia and highlight the potential of APs with pronounced anti-inflammatory action to mitigate these effects. Overall, these observations should be considered preliminary signals that warrant replication in larger prospective cohorts.

## Supporting information

STROBE check-list

## Data Availability

All data produced in the present study are available upon reasonable request to the authors

## Approval of the Ethics committee

This study was approved by the Research Ethics Board of the National Institute of Mental Health in Klecany (Approval No. 127/17).

## Declaration of generative AI and AI-assisted technologies in the writing process

During the preparation of this work, the authors used QuillBot in order to improve the readability and language of this narrative review. After using this tool/service, the authors reviewed and edited the content as needed and take full responsibility for the content of the published article.

## Contributors

**NS** performed the statistical analysis, calculated hemato-immunological ratios (NLR, MLR, PLR, and SII), computed cumulative chlorpromazine equivalent doses (cCPZ), applied the inclusion and exclusion criteria, and wrote the manuscript. **MK** and **IT** assisted with the statistical analysis and provided detailed proofreading and editorial suggestions to improve the manuscript. Additionally, **IT** prepared the STROBE checklist. **FS** conceived the original research idea, designed the overall study, supervised its implementation as head of the research group, and contributed to theoretical framing and manuscript revision. **VC** provided statistical oversight by verifying the analyses. **PF** was responsible for data extraction from the ESO database.

All authors reviewed and approved the final manuscript.

## Acknowledgements

The authors would like to thank the Psychiatric Hospital Bohnice for their collaboration and support during the initial phase of data collection. We would also like to thank the National Institute of Mental Health in Klecany for providing access to the ESO database, which was essential for conducting this study.

