## Supplemental Table 1 for "Antipsychotics Lower Peripheral Markers of Inflammation in Drug-naïve Early Psychosis: A Pilot Study"

*Bonferroni adjustment*

| **Marker** | **Raw p** | **Bonferroni p_adj** |
| --- | --- | --- |
| ΔNLR | 0.0484* | 0.1936 |
| ΔMLR | 0.0409* | 0.1636 |
| ΔPLR | 0.0277* | 0.1108 |
| ΔSII | 0.0276* | 0.1104 |

** statistical significance p>0.05*
