## Supplementary material for "Antipsychotics Lower Peripheral Markers of Inflammation in Drug-naïve Early Psychosis: A Pilot Study": STROBE check-list

STROBE Statement—Checklist of items that should be included in reports of ***cohort studies***

Shorter name: Anti-inflammatory Effect of Antipsychotics: A Pilot Study

Authors: Nicole Šafářová^1,2,3^, Marián Kolenič ^1,2^, Ivana Tašková ^3,4^, Václav Čapek ^1^, Petra Fürstová ^1^, Filip Španiel ^1,2^

Affiliations:

^1^ National Institute of Mental Health, Klecany, Czech Republic

^2^ 3rd Faculty of Medicine at Charles University, Prague, Czech Republic

^3^ Psychiatric Hospital Bohnice, Prague, Czech Republic

^4^ Faculty of Pharmacy at Charles University, Hradec Králové, Czech Republic

|  | | **Item No** | **Recommendation** | **Page No** |
| --- | --- | --- | --- | --- |
| **Title and abstract** | | 1 | (*a*) Indicate the study’s design with a commonly used term in the title or the abstract  Title – A pilot study. | 1 |
|  |  |  | (*b*) Provide in the abstract an informative and balanced summary of what was done and what was found  Monitoring inflammatory markers in FEP over time in relation to cCHPMZ; dose-dependent reductions of inflammatory markers. | 2 |
| **Introduction** | | | | |
| Background/rationale | | 2 | Explain the scientific background and rationale for the investigation being reported  Describes neuroinflammation in schizophrenia and the unclear anti-inflammatory effects of antipsychotics. | 3 |
| Objectives | | 3 | State specific objectives, including any prespecified hypotheses  Aim was to assess the association between cumulative antipsychotic exposure and inflammatory marker changes. | 3-4 |
| **Methods** | | | | |
| Study design | | 4 | Present key elements of study design early in the paper  Retrospective pilot cohort study. | 4 |
| Setting | | 5 | Describe the setting, locations, and relevant dates, including periods of recruitment, exposure, follow-up, and data collection  Psychiatric Hospital Bohnice and NIMH Klecany, Czech Republic. | 4 |
| Participants | | 6 | (*a*) Give the eligibility criteria, and the sources and methods of selection of participants. Describe methods of follow-up  Inclusion criteria (drug-naïve FEP, continuous hospitalization, no infection, monitored medication); N = 33. | 4 |
|  |  |  | (*b*) For matched studies, give matching criteria and number of exposed and unexposed  Not a matched study. | N/A |
| Variables | | 7 | Clearly define all outcomes, exposures, predictors, potential confounders, and effect modifiers. Give diagnostic criteria, if applicable Outcomes = change in NLR, MLR, PLR, SII; exposure = cCHPMZ; confounders = sex, age, DUP, clozapine use. | 4-5 |
| Data sources/ measurement | | 8* | For each variable of interest, give sources of data and details of methods of assessment (measurement). Describe comparability of assessment methods if there is more than one group  CBCs used for inflammatory markers; cCHPMZ calculated from recorded dosages; measurement formulas specified. | 4-5 |
| Bias | | 9 | Describe any efforts to address potential sources of bias  Confounding addressed through multivariate regression. | 5 |
| Study size | | 10 | Explain how the study size was arrived at  N = 33, justified as a pilot; post hoc power analysis included. | 4-5 |
| Quantitative variables | | 11 | Explain how quantitative variables were handled in the analyses. If applicable, describe which groupings were chosen and why  Quantitative variables: cCHPMZ as continuous; formulas defined for ratios; regression models used; outliers not excluded. | 4-5 |
| Statistical methods | | 12 | (*a*) Describe all statistical methods, including those used to control for confounding  Linear/multiple regression.  FDR Benjamini-Hochberg correction was used to adjust all *p*-values. | 5-6 |
|  |  |  | (*b*) Describe any methods used to examine subgroups and interactions  Subgroup (sex, clozapine) effects tested. | 5-6 |
|  |  |  | (*c*) Explain how missing data were addressed  None. | 5 |
|  |  |  | (*d*) If applicable, explain how loss to follow-up was addressed  No loss to follow-up (inpatients). | 4 |
|  |  |  | (*e*) Describe any sensitivity analyses  Sensitivity/power analysis reported. | 6 |
| **Results** | | | |  |
| Participants | | 13* | (a) Report numbers of individuals at each stage of study—eg numbers potentially eligible, examined for eligibility, confirmed eligible, included in the study, completing follow-up, and analysed  33 included; all met criteria. | 6 |
|  |  |  | (b) Give reasons for non-participation at each stage  No dropouts; reasons for non-participation not applicable. | 6 |
|  |  |  | (c) Consider use of a flow diagram  Flowchart not used due to simple inclusion. | N/A |
| Descriptive data | | 14* | (a) Give characteristics of study participants (eg demographic, clinical, social) and information on exposures and potential confounders  Age, sex, DUP, BMI, days V0-V1, baseline values. | 6 |
|  |  |  | (b) Indicate number of participants with missing data for each variable of interest  No missing data. | 6 |
|  |  |  | (c) Summarise follow-up time (eg, average and total amount)  Mean duration of follow-up: ~37 days. | 6 |
| Outcome data | | 15* | Report numbers of outcome events or summary measures over time  ΔNLR, ΔMLR, ΔPLR, ΔSII reported with summary stats and regression results. | 6-7 |
| Main results | 16 | (*a*) Give unadjusted estimates and, if applicable, confounder-adjusted estimates and their precision (eg, 95% confidence interval). Make clear which confounders were adjusted for and why they were included  Estimates with 95% CIs presented; adjusted for covariates. | | 7-10 |
|  |  | (*b*) Report category boundaries when continuous variables were categorized  No categorization of continuous variables. | | 7 |
|  |  | (*c*) If relevant, consider translating estimates of relative risk into absolute risk for a meaningful time period  Not applicable (no risk measures). | | N/A |
| Other analyses | 17 | Report other analyses done—eg analyses of subgroups and interactions, and sensitivity analyses  Sex and clozapine tested as predictors; effect sizes and multicollinearity assessed; FDR Benjamini-Hochberg correction was done. | | 10-11 |
| **Discussion** | | | | |
| Key results | 18 | Summarise key results with reference to study objectives  cCHPMZ predicted decreases in inflammatory markers; sex affected MLR. | | 7-11 |
| Limitations | 19 | Discuss limitations of the study, taking into account sources of potential bias or imprecision. Discuss both direction and magnitude of any potential bias  Small sample size, retrospective design, absence of control group. | | 11-12 |
| Interpretation | 20 | Give a cautious overall interpretation of results considering objectives, limitations, multiplicity of analyses, results from similar studies, and other relevant evidence  Results suggest potential anti-inflammatory effects of antipsychotics; calls for larger studies. | | 13 |
| Generalisability | 21 | Discuss the generalisability (external validity) of the study results  Limited; findings are preliminary, applicable to early psychosis with continuous inpatient care. | | 13 |
| **Other information** | | | | |
| Funding | 22 | Give the source of funding and the role of the funders for the present study and, if applicable, for the original study on which the present article is based  Ministry of Health CZ, grant NU22-04-00143. Johannes Amos Comenius Programme (P JAC) provided by MSMT, reg. number CZ.02.01.01/00/23_020/0008560 Role of funders not specified. | | 14 |

14

*Give information separately for exposed and unexposed groups.

**Note:** An Explanation and Elaboration article discusses each checklist item and gives methodological background and published examples of transparent reporting. The STROBE checklist is best used in conjunction with this article (freely available on the Web sites of PLoS Medicine at http://www.plosmedicine.org/, Annals of Internal Medicine at http://www.annals.org/, and Epidemiology at http://www.epidem.com/). Information on the STROBE Initiative is available at http://www.strobe-statement.org.
